# High-frequency screening combined with diagnostic testing for control of SARS-CoV-2 in high-density settings: an economic evaluation of resources allocation for public health benefit

**DOI:** 10.1101/2021.03.04.21252949

**Authors:** Will Rogers, Manuel Ruiz-Aravena, Dale Hansen, Wyatt Madden, Maureen Kessler, Matthew W. Fields, Matthew J. Ferrari, Connie B. Chang, Jayne Morrow, Andrew Hoegh, Raina K. Plowright

## Abstract

SARS-CoV-2 spreads quickly in dense populations, with serious implications for universities, workplaces, and other settings where exposure reduction practices are difficult to implement. Rapid screening has been proposed as a tool to slow the spread of the virus; however, many commonly used diagnostic tests (e.g., RT-qPCR) are expensive, difficult to deploy (e.g., require a nasopharyngeal specimen), and have extended turn-around times. We evaluated testing regimes that combined diagnostic testing using qPCR with high-frequency screening using a novel reverse-transcription loop-mediated isothermal amplification (RT-LAMP, herein LAMP) assay. We used a compartmental susceptible-exposed-infectious-recovered (SEIR) model to simulate screening of a university population. We also developed a Shiny application to allow administrators and public health professionals to develop optimal testing strategies given site-specific assumptions about testing investment, target population, and cost. The frequency of screening, especially when pooling samples, was more important for minimizing epidemic size than test sensitivity, behavioral compliance, contact tracing capacity, and time between testing and results. Our results suggest that when testing budgets are limited, it is safer and more cost-effective to allocate the majority of funds to screening. Rapid, cost-effective, and scalable screening tests, like LAMP, should be viewed as critical components of SARS-CoV-2 testing in high-density populations.

## Introduction

Vaccine coverage sufficient to reach the herd immunity threshold will take time (*10 Global Health Issues to Track in 2021*, n.d.); consequently, “test-trace-isolate” policies continue to be critical to control the spread of severe acute respiratory syndrome coronavirus (SARS-CoV-2) that causes coronavirus disease 2019 (COVID-19; Larremore et al., 2021). Even with adequate investment in human resources and infrastructure, epidemics can quickly exceed available reagents and supplies (Babiker et al., 2020) and force delays in testing turnaround and contact tracing (Clark et al., 2020; Tromberg et al., 2020). Amidst growing and urgent demands to maintain open societies, testing resources need to be allocated efficiently to optimize epidemiological outcomes.

Diagnostic testing has largely been based on reverse transcription-quantitative polymerase chain reaction (RT-qPCR); the cost of RT-qPCR testing is high, and it requires hours to perform with highly trained personnel and equipment. If testing is restricted to symptomatic individuals, many cases may be missed, particularly in universities and schools where younger individuals are more likely to be asymptomatic (Davies et al., 2020) and asymptomatic transmission of SARS-CoV-2 may comprise the majority of transmission (59%; Johansson et al., 2021). Therefore, symptom-based screening alone may not be adequate to prevent community transmission in high risk groups such as college campuses ((Paltiel et al., 2020)) and workplaces (see CDC, 2020 and Fragala et al., 2020). Additionally, RT-qPCR is expensive, and many institutions have reserved testing for symptomatic individuals (Appendix 4). The impact of focusing on only testing the symptomatic population alone may partly explain why US counties with college campuses saw coronavirus deaths double from the end of August through early December 2020, while communities without university populations only increased by half in coronavirus deaths during the same period (see Ivory et al., 2020 and Watson et al., 2020).

Alternative surveillance strategies embrace rapid tests like LAMP and point-of-care antigen-based tests to detect infectious individuals for a fraction of the cost of RT-qPCR. These tests have lower sensitivity than RT-qPCR (reviewed by Jayamohan et al., 2021). Unlike antigen-based tests, however, LAMP is sensitive enough to serve as a diagnostic tool (Nagura-Ikeda et al., 2020) when implemented in a laboratory operating under CLIA guidelines, with identification of infectious persons within 30-45 minutes. Quick turnaround facilitates rapid isolation of patients and follow on contact tracing. Moreover, the lower cost of these assays compared to PCR allows more frequent and widespread testing, creating better epidemiological outcomes (Larremore et al., 2021; Mina & Andersen, 2020). However, there has been a reluctance to use rapid, inexpensive tests (West et al., 2020) despite evidence that lower sensitivity may only be truly relevant for a short period when individuals are first infectious (Mina et al., 2020) and that high-frequency screening can compensate for the lower sensitivity (Larremore et al., 2021; Mina et al., 2020; Paltiel et al., 2020). Strategies that leverage the testing capacity of novel screening assays may yield better epidemic outcomes despite false-negative results existing. However, it is unclear how to optimally invest limited funds between diagnostic and screening testing to minimize epidemic size in resource-limited testing scenarios (Vandenberg et al., 2021).

Here, we simulated the widespread and frequent screening of students on a university campus to assess the optimal magnitude and frequency of testing required to control an outbreak of SARS-CoV-2 during a 15-week semester. We explored the implementation of an inexpensive, rapid, and scalable SARS-CoV-2 test based on loop-mediated isothermal amplification (LAMP) technology on a hypothetical university campus (20,000 students) mixed with diagnostic RT-qPCR testing. We approached modeling with three main goals: (Objective 1) to evaluate the epidemiological effects of screening; (Objective 2) to determine the effect of assay sensitivity, student behavior, screening frequency, and sample pooling approaches on epidemiological outcomes; (Objective 3) to determine the optimal ratio of diagnostic testing to screening testing for resource allocation optimization. Finally, we implement our model framework in an interactive web application, created with the Shiny R package, where decision-makers can compare the epidemiological and demographic context of respective communities with testing capacities and calculate the level of screening required to reach desired targets of epidemic control.

## Methods

### Objective 1: Screening capacity to limit epidemics and moderators of screening efficacy

We simulated testing programs with screening coverage from 0% to 20% of the population per day and test sensitivity ranging from 60-90%, reflecting the range of sensitivities of alternative tests to RT-qPCR (Mina & Andersen, 2020). We recorded total epidemic size in a 15 week period (reported as a percent of the total population), quantity of diagnostic tests needed to test symptomatic individuals (unadjusted for positivity; i.e., additional tests may be needed for people with non-COVID, influenza-like illness), quantity of RT-PCR assays needed to confirm cases detected by screening, and daily prevalence of active infections. Symptomatic tests were assumed to be unlimited in this simulation - diagnostic-limited scenarios under objective 3. We also evaluated how population compliance (50% or 100% compliance), the sensitivity of the screening assay (60-90%), and the screening frequency (0-20% of the population per day) affected the screening efficacy. We quantified efficacy through total epidemic size as well as through the proportion of active cases discovered by a testing scenario relative to the total active infection days in an epidemic.

### Objective 2: Implementing screening

We then investigated how effectively screening helped control the epidemic by varying the number of tests deployed over time and by pooling samples for testing. Four implementation strategies were considered with varying distributions of tests over time: (1) a “symptomatic only” scenario with no asymptomatic screening; (2) an “initial effort” strategy which focused all screening testing effort on the first 30 days of the semester, with no screening from day 31 through 150; (3) a “front-loaded” scenario which split all screening tests equally between the first 30 days of the semester and the last 120; and (4) a sustained effort scenario which evenly dispersed screening assays over the semester. Sample pooling can increase testing capacity, especially when the prevalence of active infections in the population is low (Mutesa et al., 2021). However, pooling can introduce error into testing (Heffernan et al., 2014) and dilution of samples can decrease cycle threshold values (Das et al., 2020). The effect of pooling likely varies across assays used and implementation, but the loss of sensitivity is generally considered to be minor (Barat et al., 2020; Das et al., 2020; Perchetti et al., 2020). We assumed that pooling acted as a multiplier on the daily number of tests available with a 5% penalty in sensitivity for each additional pooling factor (*e*.*g*., if a single-replicate test is 77% sensitive, a two-sample pool is 72% sensitive). Scenarios of test capacity without pooling ranged from 2.5%-20% of the population per day (500 to 4000 tests available per day). Pools of two and four samples per test increased test capacity to 5-40% and 10-80% of the population per day, respectively. Screening assay sensitivity without pooling was assumed to be 77%, based on estimation from field deployment of LAMP (Chang et al. in prep).

### Objective 3: Optimum combination of screening and diagnostic testing

We evaluated how to optimally distribute funding between diagnostic testing and screening for the best epidemiological outcomes. The basic SEIR model was modified so that a defined budget was allocated equally over time with some proportion towards diagnostic testing costs and the remainder towards screening costs. Inclusive of all costs from sample collection to the communication of results at our institution, we estimated that screening cost ∼USD$3.50 per assay (C. Chang et al., in prep), while diagnostic RT-qPCR cost ∼USD$12.50 per assay (S. Walk, personal comm.). In general, assays useful in screening programs are at least 2-10 times cheaper than RT-qPCR (see Ravi et al., 2020). If there were fewer symptomatic cases arriving at care facilities than diagnostic RT-qPCR tests available, excess tests were rolled-over into asymptomatic testing, providing a conservative comparison between diagnostic testing and screening. The scenarios included testing budgets from USD$0 to USD$2 million (USD$0 to USD$0.67 per individual per day), allowing the investment of funds towards screening (and consequently divestment from diagnostic testing) to range from 0% to 100%. We also considered individual compliance and test sensitivity as moderators of the screening scenarios. We used two proxies to assess the efficiency of the testing programs: the total epidemic size, and the total cost (inclusive of all diagnostic, screening, and confirmatory tests as well as isolation and quarantine costs).

This analysis adheres to the Consolidated Health Economic Evaluation Reporting Standards ([CHEERS](https://www.equator-network.org/reporting-guidelines/cheers/)) reporting guidelines, where applicable.

## Results

### Objective 1: Screening capacity to limit epidemics and moderators of screening efficacy

Testing frequency had a greater effect on epidemic size than sensitivity (Fig. 2:A; Appendix 3 SI: Fig. 1), individual compliance (Fig. 2:A), and delays in testing turn-around time (Appendix 3 SI: Fig. 1) or limits on contact tracing (Appendix 3 SI: Fig. 3). Higher screening frequencies led to the discovery of more infectious cases such that the higher the proportion tested, the lower the epidemic size (Fig. 2; Appendix 3 SI: Fig. 2). Higher screening frequencies decreased the demand for diagnostic RT-qPCR tests (Fig. 2:C) but increased discovery of cases initially led to the need for more confirmatory RT-qPCR tests (Fig. 2:D). Screening frequencies greater than 10-20% sufficiently reduced the epidemic to compensate for higher discovery rate of infections (Fig. 2:D). Compliance was equally or potentially more influential on epidemic size than screening sensitivity, depending upon the frequency of screening. Increasing individual compliance from 50% to 100% decreased epidemic size by as much as 24.6% (assuming 10% of the population tested per day and 75% screening sensitivity) - roughly equivalent to testing an additional 4.5% of the population per day with 100% compliance. Individual compliance also affected the proportion of infectious cases detected; with 100% compliance, testing programs detected up to 9.4% more cases than programs with only 50% compliance (with 90% screening sensitivity and 20% of the population tested per day).

**Figure 1:**
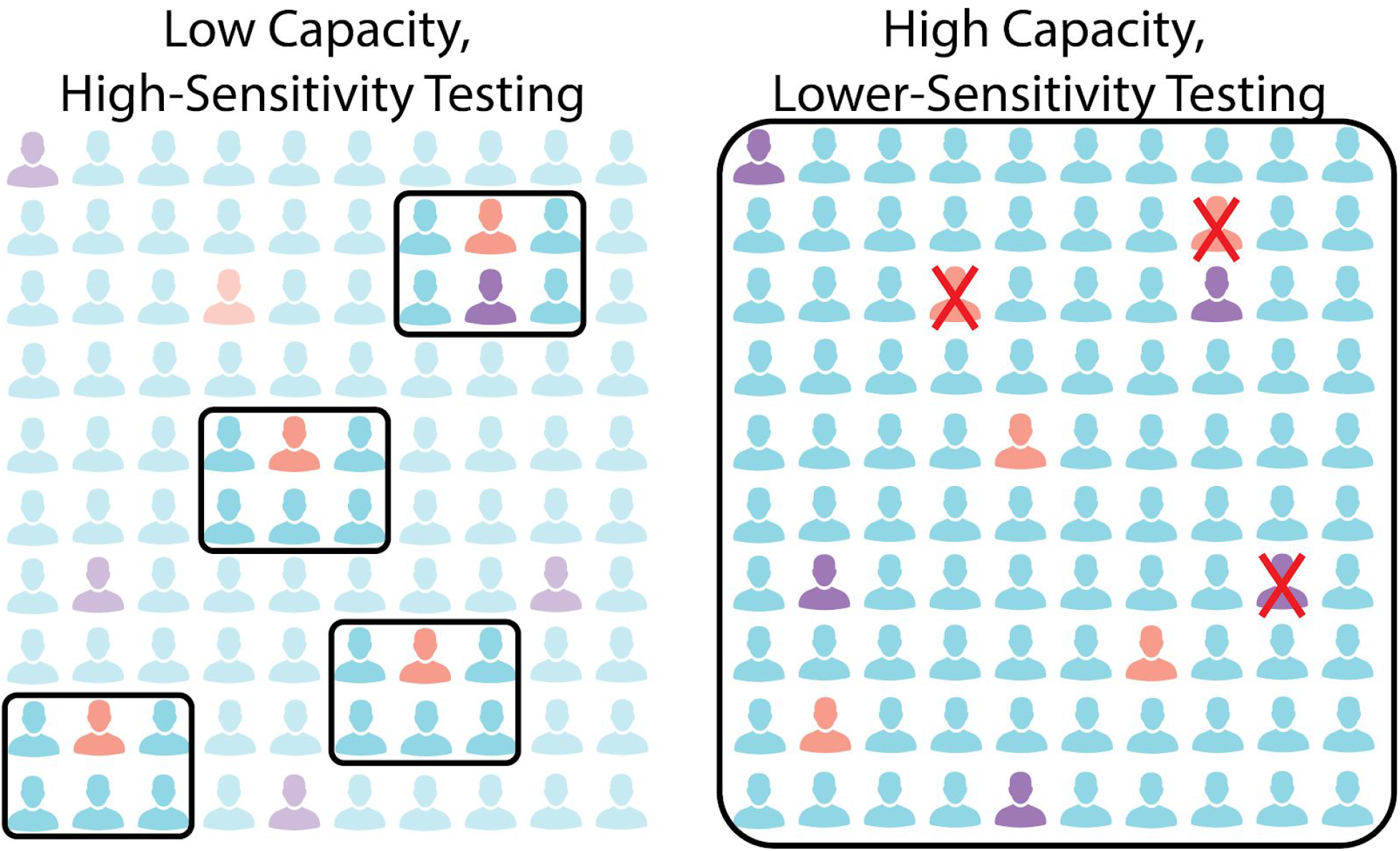
Infected individuals who are symptomatic (red, 5%) and asymptomatic (purple, 5%) along with healthy individuals (blue, 90%) comprise a theoretical population. Untested individuals are more transparent, and individuals with red crosses are false-negative results. At left, diagnostic tests are used to test symptomatic individuals and quarantine their contacts; five infectious individuals are removed from the population (four symptomatic and one asymptomatic contact). At right, screening (70% sensitive) is used to test the population randomly; seven infectious individuals are removed from the population (three symptomatic and four asymptomatic cases) even though three individuals were misdiagnosed.

**Figure 2:**
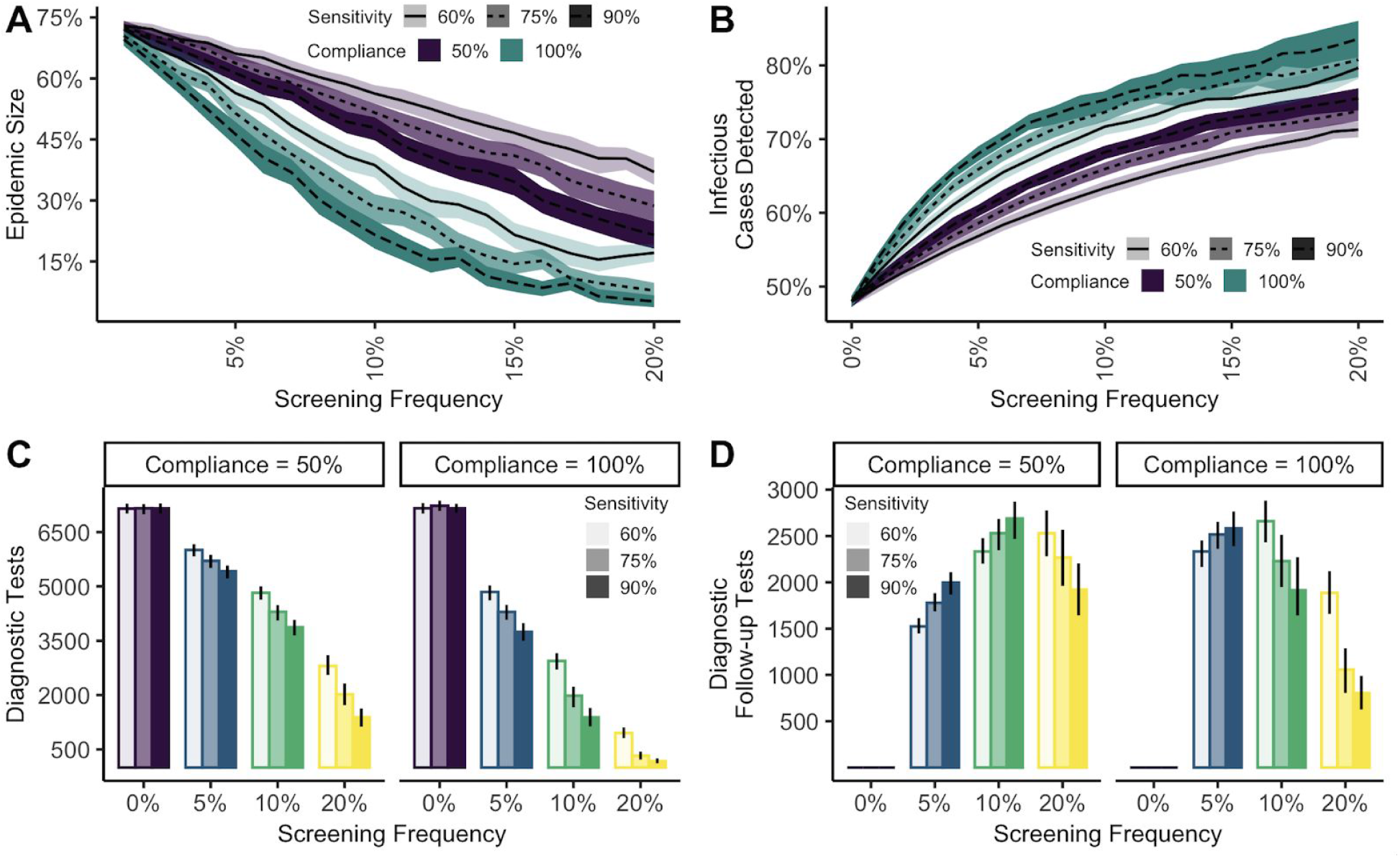
The effect of testing strategies on (A) epidemic size stratified by compliance (color of band) and sensitivity (opacity and line type); (B) the proportion of infectious cases detected by diagnostic and screening testing stratified by compliance (color of band) and sensitivity (opacity and line type); (C) symptomatic RT-qPCR positive tests determined by compliance (facet), sensitivity (opacity), and testing frequency (color); and, (D) screening follow-up confirmatory RT-qPCR tests determined by compliance (facet), sensitivity (opacity), and testing frequency (color). Error bars and ribbons indicate the 95% quantile of observation from 200 simulations.

### Objective 2: Implementing screening

We considered four scenarios of how universities, businesses, or other dense populations might structure testing for a general population considering the allocation of tests in time, capacity, and pooling factors (“symptomatic-only”, “initial effort”, “sustained effort”, and “front-loaded effort”, from Methods). The distribution of tests over time and the pooling factor were critical to the overall epidemic size and shape (Fig. 3; Appendix 3 SI: Fig. 5). In the most restricted testing scenario with only 500 tests available per day and no pooling of screening samples, sustained testing led to the greatest difference in epidemic size as compared to a symptomatic-only testing scenario (11%), followed by front-loaded testing (8.4%), and initial effort testing (2.6%). With greater pooling factors and more tests available per day, front-loaded testing scenarios led to smaller epidemics than sustained testing scenarios. Initial effort scenarios were generally only successful at reducing epidemic size at higher testing frequencies and with higher pooling factors (Fig. 3) and led to epidemics that peaked later in the semester (Appendix 3 SI: Fig. 5). Despite a conservative 5% penalty in sensitivity with each additional pool, pooling factors of 2 and 4 always produced lower epidemics than no-pooling scenarios.

**Figure 3:**
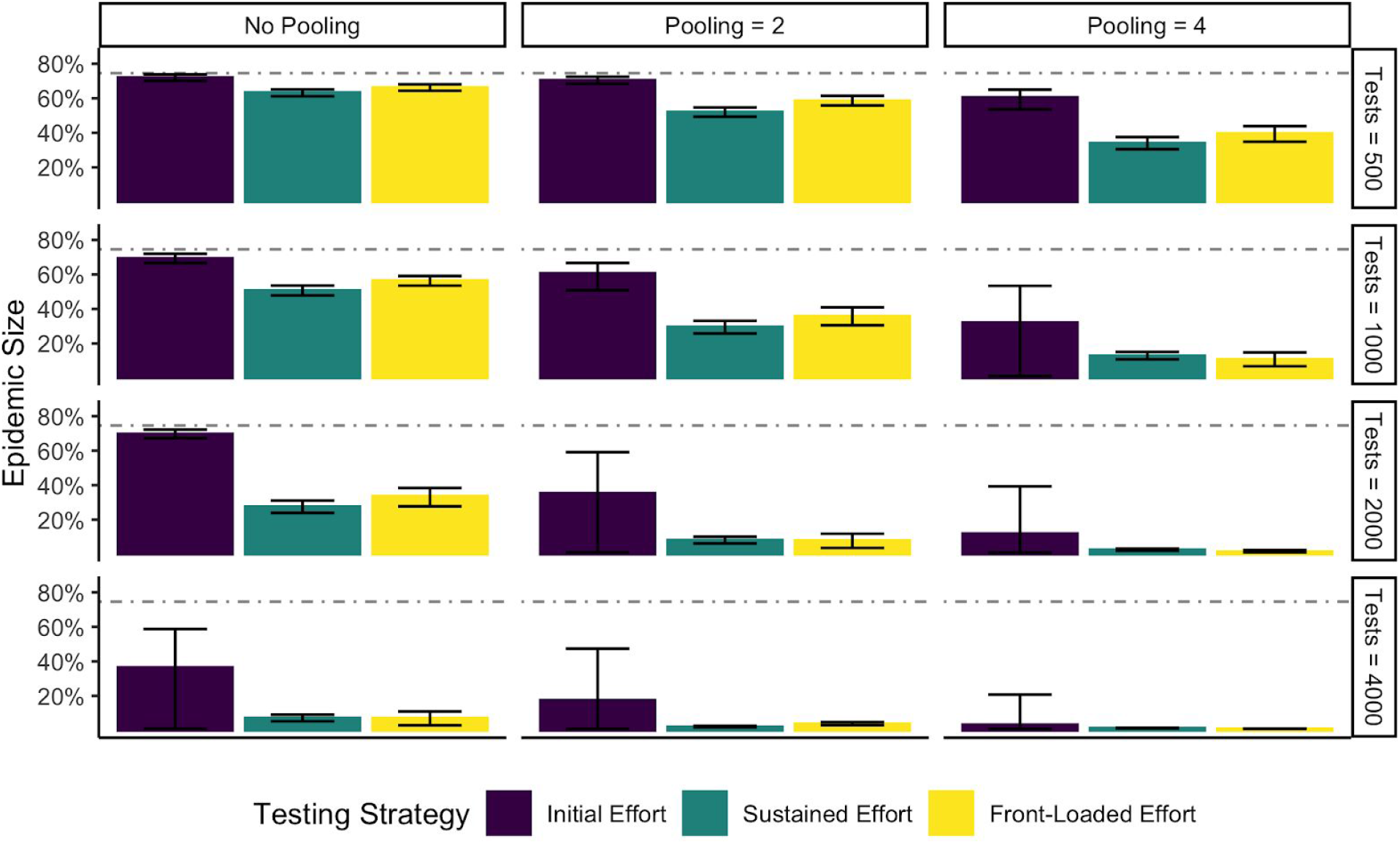
The epidemic size of “initial”, “sustained”, and “front-loaded” LAMP testing scenarios (from Methods) faceted by pooling factors (one, two, and four samples per test) and by the number of available tests per day (500-4000 tests, or 2.5-20% of the population). Colored bars represent the mean epidemic size and error bars represent the 95% quantile across 200 simulations at each combination of variables. The dashed grey line denotes the mean epidemic size associated with symptomatic-only testing scenarios.

### Objective 3: Optimum combination of screening and diagnostic testing

Finally, we evaluated how testing funds can be distributed between screening and diagnostic testing to have the greatest impact on epidemic size. Total cost was partially structured by investment in testing, ranging from $0 to $2 million USD (USD$0 to USD$100 per individual per semester), but also considered the cost of confirmation testing (same as diagnostic testing; USD$12.50) and isolation and quarantine of individuals who lived on campus (USD$25 per individual per day), a conservative estimate of potential costs to a university. For every combination of investment in screening and diagnostic testing, individual compliance, and screening test sensitivity, we reported the percent investment in screening that returned the lowest epidemic size (Fig. 4); a full comparison is provided in Appendix 3 SI: Fig. 6). As the available budget for testing increased, the proportion of funds devoted to screening increased in order to minimize epidemic size (Fig. 4); however, as screening costs increase, more investment should be allocated to diagnostic costs (Appendix 3 SI: Fig. 7:A). If, for example, a university had USD$0.2 mil (USD$10 per student per semester) to invest in testing (both diagnostic and screening), epidemic sizes are minimized when all testing funds are invested in diagnostic testing. If a university can expend USD$2 mil (USD$100 per student per semester) on testing, the university should invest upwards of 95% of funds towards screening to reduce epidemic size. Scenarios with higher-sensitivity screening assays, greater compliance, and pooled testing had a higher optimal proportion of investment in screening. For instance, scenarios with 90% sensitivity, had optimal investment proportions 0.025 (sd=0.038) greater than scenarios with 60% sensitivity; scenarios with 100% compliance had optimal investment proportions 0.048 (sd = 0.46) greater than scenarios with 50% compliance; and finally, scenarios with a pooling factor of four had optimal investment proportions 0.098 (sd = 0.94) greater than scenarios with no pooling.

**Figure 4:**
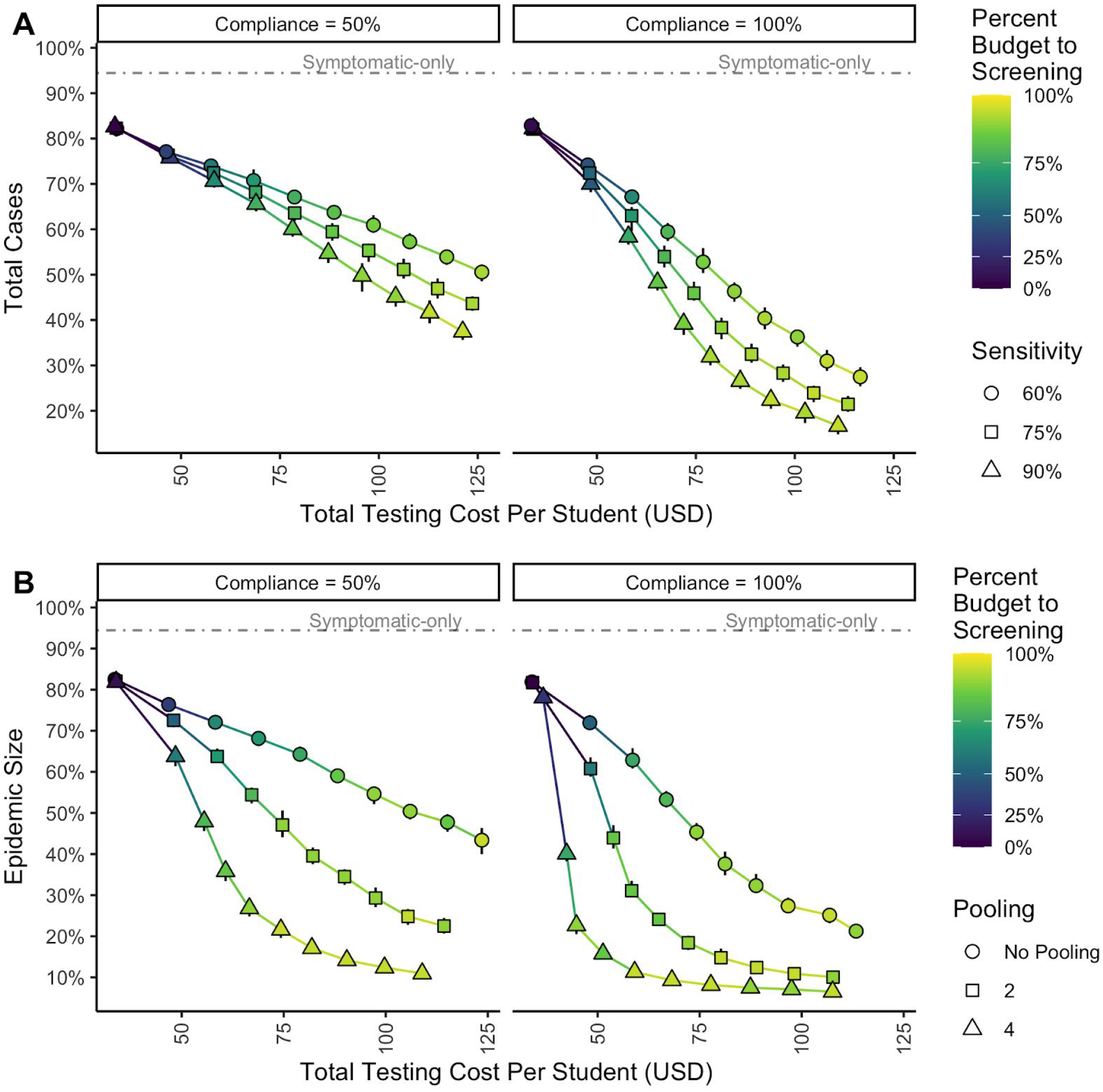
The relationship between monetary investment in testing (diagnostic and screening) and epidemic size. Points represent the mean cost and mean cases of the smallest epidemic scenario at every combination of financial investment in testing. The shape of each point corresponds to (A) the sensitivity underlying a given scenario or (B) the pooling factor underlying a given scenario. The color of each point is the proportion of money invested in screening to minimize epidemic size at a given combination of compliance (facet), testing investment, sensitivity, and pooling factor. Error bars represent the 95% quantile across 25 simulations at each combination of variables. The dashed grey line denotes the mean epidemic size associated with symptomatic-only testing scenarios.

**Figure 5:**
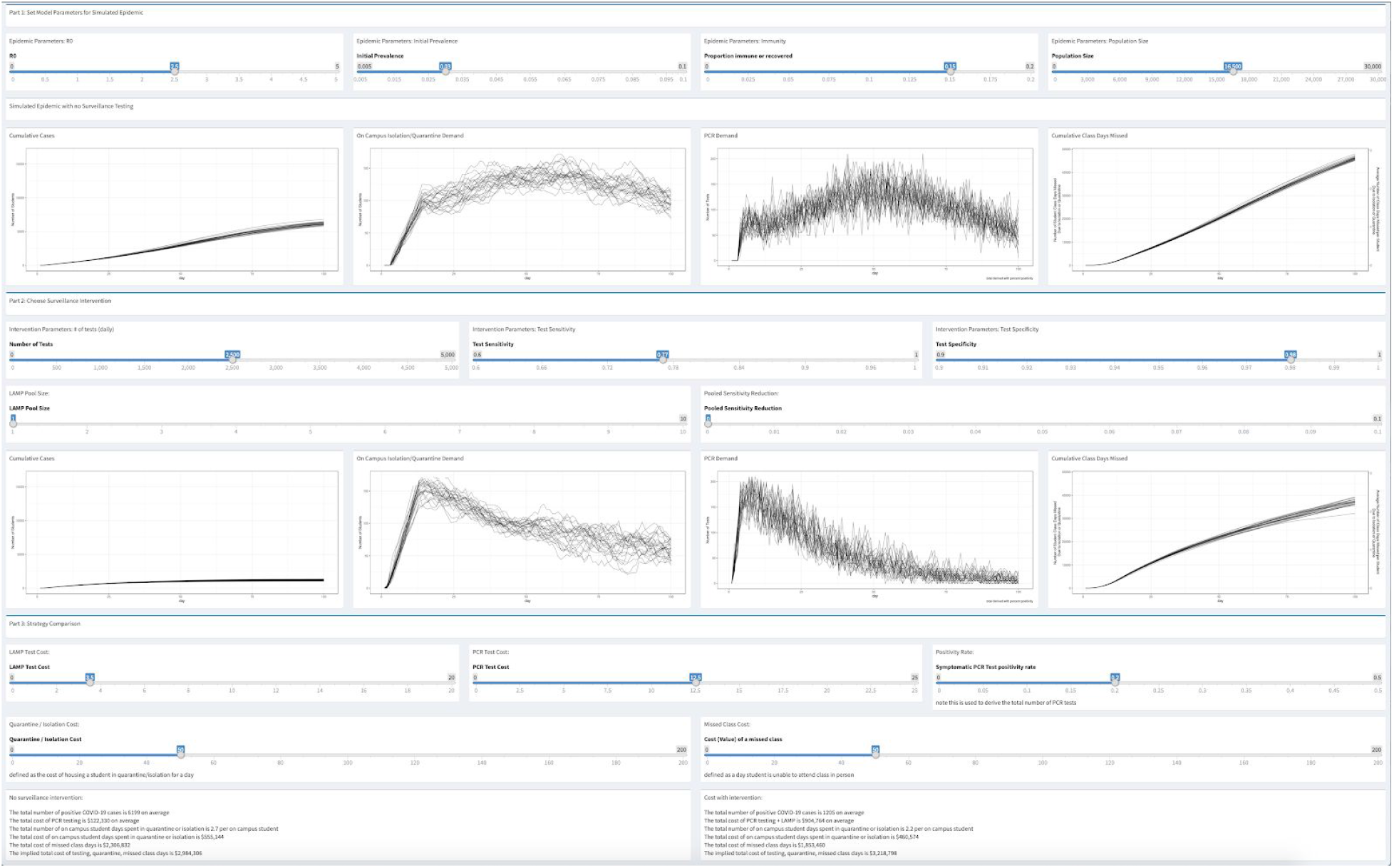
A complete view of the Shiny application using our model under a variety of epidemic conditions, comparing symptomatic only testing to screening scenarios with output about the epidemic size, isolation and quarantine capacity, demand on point of care facilities, effects on student-days in class, and the cost of testing scenarios.

### Implementation of tool for decision-making

We built a Shiny app to allow comparison of multiple testing strategies with user-specific epidemiological and demographic assumptions about the underlying population, testing capacity, and the cost structure of testing. The module reports epidemic size, estimates of isolation and quarantine occupancy, demand on testing and point-of-care facilities, and the number of student-days missed. While the total cost for screening scenarios is generally higher than symptomatic-only testing scenarios, this app is useful for showing how assay capacity and pooling can generate safer and more cost-effective epidemic containment strategies in simulated university conditions. The Shiny app and the toggles available to the user are discussed in Appendix 2.

## Discussion

Non-pharmaceutical interventions such as isolation of infected individuals, social distancing, and face mask use will limit harm and loss of life from COVID-19 until vaccinated and recovered individuals provide herd immunity (Gozzi et al., 2021). Complementing these measures with frequent testing of the population and effective post-testing behavioral controls will be critical to maintain in-person function of schools, universities, and other high-density populations (Paltiel et al., 2020). Our results showed that lower-sensitivity but higher-frequency tests can be a valuable tool to limit SARS-CoV-2 epidemics, particularly when used in conjunction with diagnostic testing and sample pooling. While we focused on testing programs utilizing LAMP, our work applies to other rapid and inexpensive tests, such as antigen-based testing, which have lower sensitivity than RT-qPCR but comparable turn-around speed and resource requirements to LAMP. Our findings support the implementation of mixed diagnostic and screening strategies alongside strong public messaging and information campaigns that will increase compliance with post-testing behavioral controls to optimize epidemic control in high-density populations with considerable asymptomatic transmission.

To our knowledge, no former studies have modeled optimal allocation of funding to screening and diagnostic programs. We demonstrated that epidemics were best limited through mixed testing programs with 5-20% of testing investment devoted to diagnostic RT-qPCR testing of symptomatic cases and 80-95% allocated to screening the population using LAMP. For instance, in simulations with testing investment greater than USD$0.4 mil or USD$20 per individual per semester, mixed testing programs with funds biased towards screening generated smaller epidemics than testing programs that only invested in diagnostic testing or those that only invested in screening. We reached similar conclusions about the effect of testing delays, testing frequency, and testing sensitivity as other authors (see Larremore et al., 2021), and we found pooling to be an effective mechanism to increase testing frequency and thus limit epidemics (also shown by Deckert et al., 2020, Denny et al., 2020, Mutesa et al., 2021, and Perchetti et al., 2020).

The standard dogma of SARS-CoV-2 diagnostic testing is that assays need to be sensitive to hold utility, and understandably so as false-negative results have negative consequences (Brooks & Das, 2020; West et al., 2020). However, Mina et al., (2020) posit that diagnostic and screening tests need to be evaluated by separate criteria. Unlike diagnostic tests, the effectiveness of a screening program is determined by the frequency of testing, not the sensitivity of the assay (Larremore et al., 2021; Paltiel et al., 2020). While low-sensitivity screening programs will misdiagnose some infected individuals, repeated testing within infectious periods identifies more cases overall and is better at limiting infections in the broader population than less-frequent testing with a more sensitive test.

The rapid spread of SARS-CoV-2 globally has favored the emergence of new variants, presenting worrying complications for public health (Lauring & Hodcroft, 2021). As epidemics proceed, strains will continue to evolve, threatening vaccine efficacy (*In the COVID-19 Vaccine Race, We Either Win Together or Lose Together*, n.d.). In the US, we are still months and possibly years away from reaching levels of immunization to halt the spread of the virus (“More Than 168 Million Shots Given,” n.d.) and slow down the evolutionary capacity of SARS-CoV-2 to escape natural- or vaccine-derived immunity. Middle- and low-income countries have been particularly hard-hit by vaccine nationalism and distribution inequity, with limited testing and health care capacity to handle epidemics. High-frequency screening programs based on assays like LAMP are inexpensive, require minimal investment in materials and personnel, can readily adapt to genetic variants of SARS-CoV-2, and will work synergistically with diagnostic testing already in place. Universities, schools, businesses, and other high-density populations should reevaluate the cost of diagnostic testing, consider inexpensive and rapid alternatives like LAMP, and invest adequately in mixed screening and diagnostic programs to minimize harm and loss of life.

## Supporting information

Supplemental Information for Main Text

## Data Availability

There are no original data in this modeling study. All code to generate data used in the study are available at https://github.com/wilrogers/COVID-Modeling.

