## Supplemental Information for Main Text for "High-frequency screening combined with diagnostic testing for control of SARS-CoV-2 in high-density settings: an economic evaluation of resources allocation for public health benefit"

### Appendix 1: Model methods

#### *Model description*

We developed a campus-level, SEIR model with behavioral, testing, and transmission controls to address the implementation of loop-mediated isothermal amplification (LAMP), a generally high specificity but lower sensitivity (dependent upon replicates), rapid test for SARS-CoV-2 compared to the higher sensitivity and expensive tests, like RT-qPCR. SEIR refers to four disease compartments: susceptible, exposed (infected but not infectious), infectious, and recovered. The progression from S to E was stochastic and determined by the probability of infection (Appendix 1: EQ 1 and 2). Infectious individuals progressed from I1 (infectious, but not yet symptomatic) to I2 (infectious with both symptomatic and asymptomatic cases). The rate of progression from E to I1 was stochastic, determined by a mean incubation period of 5 days (Lauer et al., 2020; Appendix 1: EQ 3). Individuals stochastically transition from I1 to I2 based on a mean period of 2 days, (He et al., 2020; Appendix 1: EQ 4). Recognizing that a significant portion of the transmission of SARS-CoV-2 can occur before symptoms (44%; He et al., 2020), we built a boxcar compartment at I2, forcing a one-day delay until an individual could be recognized as symptomatic. After exiting the boxcar, 50% of those infectious cases were distinguished as symptomatic (Denny et al., 2020; Zhang et al., 2020). Transitions from I2 to R were determined by a post-symptomatic period of 7 days (He et al., 2020; Appendix 1: EQ 5).

$$\text{EQ 1: } S_{t,i+1} = S_{t,i}\left(e^{\frac{-\beta I_t}{N_{\mathrm{t}}}}\right)$$

$$\text{EQ 2: } E_{t,i+1} = S_{t,i}\left(1 - e^{\frac{-\beta I_t}{N_{\mathrm{t}}}}\right) - E_{t,i}\left(1 - e^{-\frac{1}{5}}\right) + \pi \mathrm{C}$$

$$\text{EQ 3: } I1_{t,i+1} = E_{t,i}\left(1 - e^{-\frac{1}{5}}\right) - I1_{t,i}\left(1 - e^{-\frac{1}{2}}\right)$$

$$\text{EQ 4: } I2_{t,i+1} = I1_{t,i}\left(1 - e^{-\frac{1}{2}}\right) - I1_{t,i}\left(1 - e^{-\frac{1}{7}}\right)$$

$$\text{EQ 5: } R_{t,i+1} = I1_{t,i}\left(1 - e^{-\frac{1}{7}}\right) + R_{t,i}$$

where S, E, I1, I2, and R represent the epidemic categories with timestep denoted by t, on- or off-campus affiliation denoted by i, and pi denotes the daily probability of community infectious introductions to the population of size C.

We included two basic subpopulation components in the model for on-campus and off-campus students. The abundance of each subpopulation was determined by enrollment data from a local university (Montana State University, Bozeman, Montana). The two subpopulations mixed homogeneously and epidemiological conditions can be modified to suit assumptions about student behavior and dormitory conditions. Individuals were removed from the mixed population in three ways: (1) symptomatic cases (a subset of I2) could elect for diagnostic testing, (2) random individuals from all compartments could be tested via random screening and removed if test-positive, (3) and random individuals from all compartments could be removed via contact tracing. We included two parameters in the model to describe how population behavior might alter the efficacy of testing and testing demand: (1) “compliance” moderated the proportion of students who participated in screening testing, who tested positive through screening, and close contacts who elected to isolate or quarantine and (2) “care-seeking” moderated the proportion of students who elected to seek diagnostic testing after demonstrating symptoms. The ability to identify symptomatic infectious cases was determined by the proportion of I2 cases that were symptomatic (50% in below simulations), the sensitivity of the test applied (diagnostic RT-qPCR is generally high, modeled at 98% below), and the probability that a student seeks care after developing symptoms (generally assumed 100% in lower simulations, but see Appendix 3 SI: Fig. 4). Symptomatic demand for tests was determined as the number of individuals in the I2 compartment who appeared at the rate of symptoms in infected individuals (50%) and the rate of care-seeking (100% here). The ability to identify infectious individuals through screening was determined by the frequency of testing (limited by daily screening limits), the sensitivity of the screening test, the compliance of students with screening testing (generally assumed 100% in below simulations unless specified), and prevalence of infection over time. Finally, infected cases were assumed to have contacts and these contacts were assigned via a random draw from a Poisson distribution with a mean of 5 (Prem et al., 2017); however, contact tracing limits constrained the number of contacts available to isolate per day. Screened individuals and contacts of test-positive individuals were removed from the mixing population by the rate of compliance, reducing the number of isolated and quarantined individuals as compliance decreased from 100%.

The length of the simulations was 150 days. $R_0$ was assumed to be 3, a rather fast-paced epidemic but not unreasonable given the epidemic trajectory on high-density populations (Salje et al., 2020). $R_{effective}$ was assumed to be $R_0$ multiplied by the proportion of non-immune individuals. $\boldsymbol{\beta}$ was assumed to be the $R_{effective}$ distributed over a nine-day infectious period. Immunity was assumed to be 5% which is reasonable based on disease incidence data from our institution at the time of writing (Healthy Gallatin, n.d.), though immunity will continue to increase with vaccination and pathogen spread (we vary immunity in Appendix 3 SI: Fig. 2). Stochastic introductions of cases to resemble community-to-campus transmission were assumed as one case every 10 days to both on-campus and off-campus subpopulations. The total population size was 20,000 with 25% of students in the on-campus subpopulation (relevant if epidemiological assumptions about on- and off-campus populations differ). Contact tracing was not considered in the body of the paper, providing a very conservative comparison of screening testing strategies to symptomatic-only testing strategies (as contact tracing increases, the efficacy of screening testing grows, relative to symptomatic-only testing). We do, however, show the effect of added contact tracing capacity in Appendix 3 SI: Fig. 3. We assumed that 50-100% of the population complies with behavioral controls post-testing (compliance), while 100% of students with symptoms seek care (care-seeking). We show how assumptions about compliance and care-seeking affect epidemic size in Appendix 3 SI: 4. Finally, within our model, we assumed that positive cases were isolated for 10 days. In general, 95% quantiles were reported for model output and diagnostics based on between 25-200 repetitions (specified in figure legends). As diagnostics of our model, we present how moderating model inputs ($R_0$, initial prevalence, initial immunity, community case introduction, contact tracing limitations, student compliance, student care-seeking, testing sensitivity, and testing strategies) within reasonable parameter space moderate total case count in simulations (Appendix 3 SI: Fig. 1-7).

All code and simulations were performed in R version 3.6.3 (*R: A Language and Environment for Statistical Computing.*, 2020) and all model scripts and associated code are available at https://github.com/wilrogers/COVID-Modeling.

Table 1: Model parameters

| Parameter | Value | Source |
|---|---|---|
| LAMP Sensitivity | 77% | Chang et al. (in prep) |
| LAMP Specificity | 98% | Chang et al. (in prep) |
| RT-qPCR Sensitivity | 98% | Wang et al., 2020 |
| RT-qPCR Specificity | 98% | Wang et al., 2020 |
| Incubation period | 5 days | Lauer et al., 2020 |
| Time to symptom onset | 2 days | He et al., 2020 |
| Percent symptomatic | 50% | Zhang et al., 2020 |
| Time to recovery | 7 days | He et al., 2020 |
| Initial prevalence | 1% | Local University |
| Initial immunity | 5% | Local University |
| $R_0$ | 3 | Salje et al., 2020 |
| Percent on-campus | 25% | Local University |
| Daily probability of community introduction on- and off-campus | 10% | Assumed |
| Size of community introduction on- and off-campus | 1 | Assumed |

### Appendix 2: Shiny app tutorial

The Shiny app for simulating an epidemic and evaluating testing interventions contains three distinct parts. The first component focuses on parameters for simulating the epidemic, without new, targeted interventions. This captures the baseline outcome at a particular location or institution. The second component of the Shiny app allows researchers to specify parameters for an intervention; in particular, sensitivity and specificity of the test, along with testing frequency can be manipulated. The final component of the model assesses the impact of the intervention by comparing public health metrics and monetizing economic costs for current strategies relative to various interventions. Given the uncertainty in model parameters and differences across locations, the Shiny app provides a user-friendly interface for researchers and decision makers to evaluate intervention strategies.

**Part 1: Setting Model Parameters for Simulated Epidemic**

There are four model parameters that need to be set to capture likely outcomes of an ongoing epidemic. $R_0$, initial prevalence, and proportion of the population that is vaccinated, recovered, or otherwise immune all control the dynamics of the epidemic. The final parameter is population size, which scales the epidemic to match the population of interest. Given these model parameters, the Shiny app generates four graphics that capture the cumulative number of positive cases as well as other metrics: quarantine/isolation housing demand, PCR testing demand, and cumulative class days missed. A snapshot of the default values can be seen in Appendix 1 SI: Fig. 1.

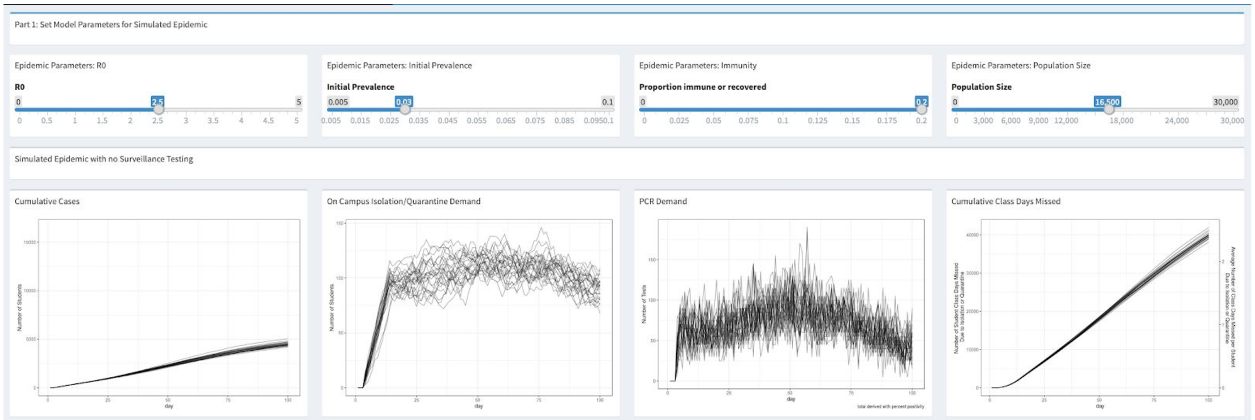

**SI Figure 1:** Part 1 of the Shiny application allows users to set parameters for the epidemic and visualize potential outcomes under those settings. The first panel contains cumulative positive cases, the second panel shows the demand for quarantine or isolation housing, the third panel contains the symptomatic PCR testing demand, and the fourth panel shows cumulative class days missed by students in isolation or quarantine.

**Part 2: Choose Surveillance Intervention**

The second component of the shiny app focuses on setting a testing intervention strategy. Users can select a daily frequency of testing along with setting test specificity and sensitivity. The Shiny app also permits pooled testing, so users can assess the impact of pooled testing strategies and potentially account for reduced test sensitivity as a function of pool size. Using the testing intervention, the Shiny app generates the same four graphics as Part 1. A snapshot of the second part of the Shiny app can be seen in Appendix 1 SI: Fig. 2.

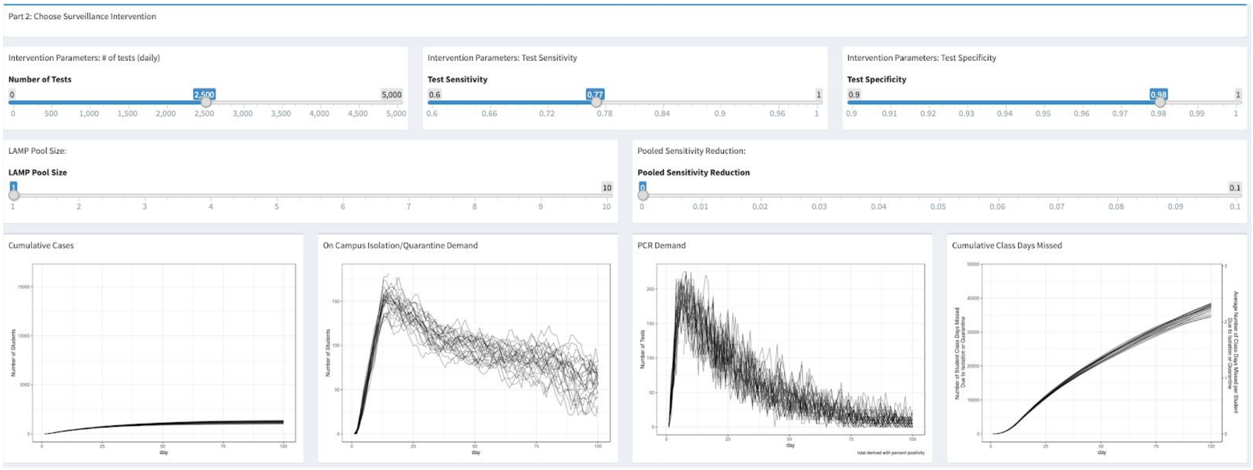

**SI Figure 2:** Part 2 of the Shiny application allows users to set testing intervention parameters for the epidemic and assess the likely outcomes. As in Figure A1.1, the output contains cumulative positive cases, demand for quarantine or isolation housing, symptomatic PCR testing demand, and cumulative class days missed by students in isolation or quarantine.

**Part 3: Choose Surveillance Intervention**

The final piece of the Shiny app summarizes the total epidemic size with and without a testing intervention. In addition to public health metrics, the app also allows economic considerations of a testing intervention. Users can enter the cost of a screening test as well as the cost of symptomatic testing.

Furthermore, economic costs can be associated with quarantine and isolation housing as well as missed class time. Finally, the app reports estimated costs associated with the status quo and a testing intervention.

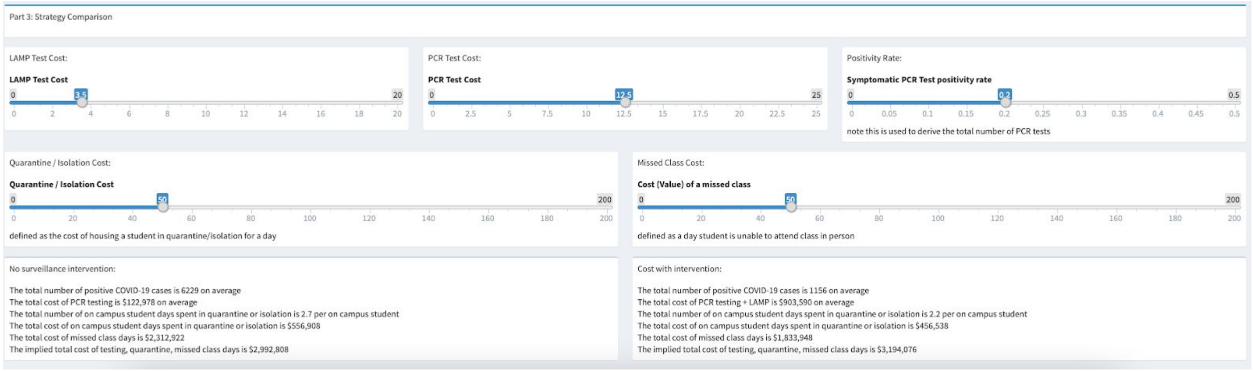

**SI Figure 3:** Part 3 of the Shiny application allows users to set economic costs associated with testing, isolation, and missed class time. This part of the shiny app also summarizes and compares expected costs between the status quo and a testing intervention.

### Appendix 3: Model sensitivity to input

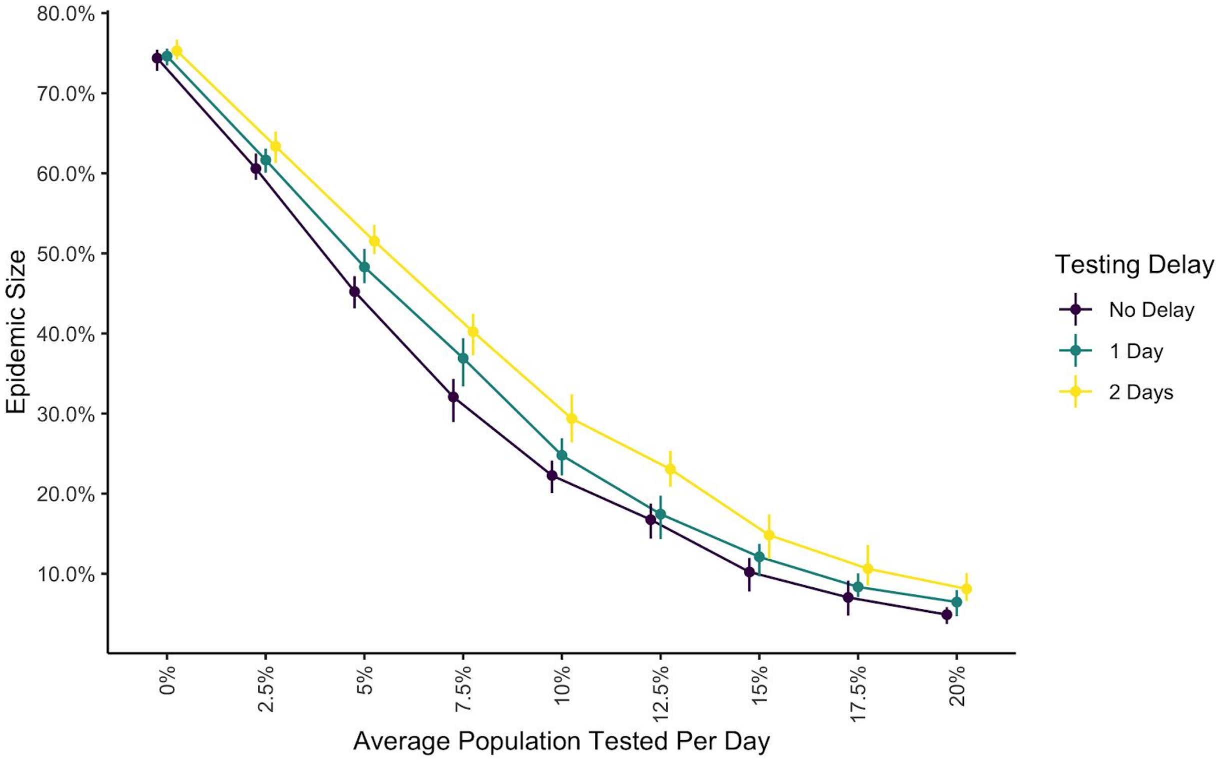

**SI Figure 1:** Epidemic size as a result of frequency of screening testing within a population of 20,000 (0-20%) and across three levels of testing delays ("no delay", "1 day", and "2 days"). This simulation used the constant testing timeline, with 100% compliance and care-seeking, 1% initial prevalence, 5% immunity, 0.5 symptomatic rate, $R_0$ of 3, PCR sensitivity and specificity of 99%, LAMP sensitivity of 77.1%, LAMP specificity of 98%, one community introduction per 10 days to both on- and off-campus, no contact tracing, no pooling, and a 150 day semester with 50 simulations per combination.

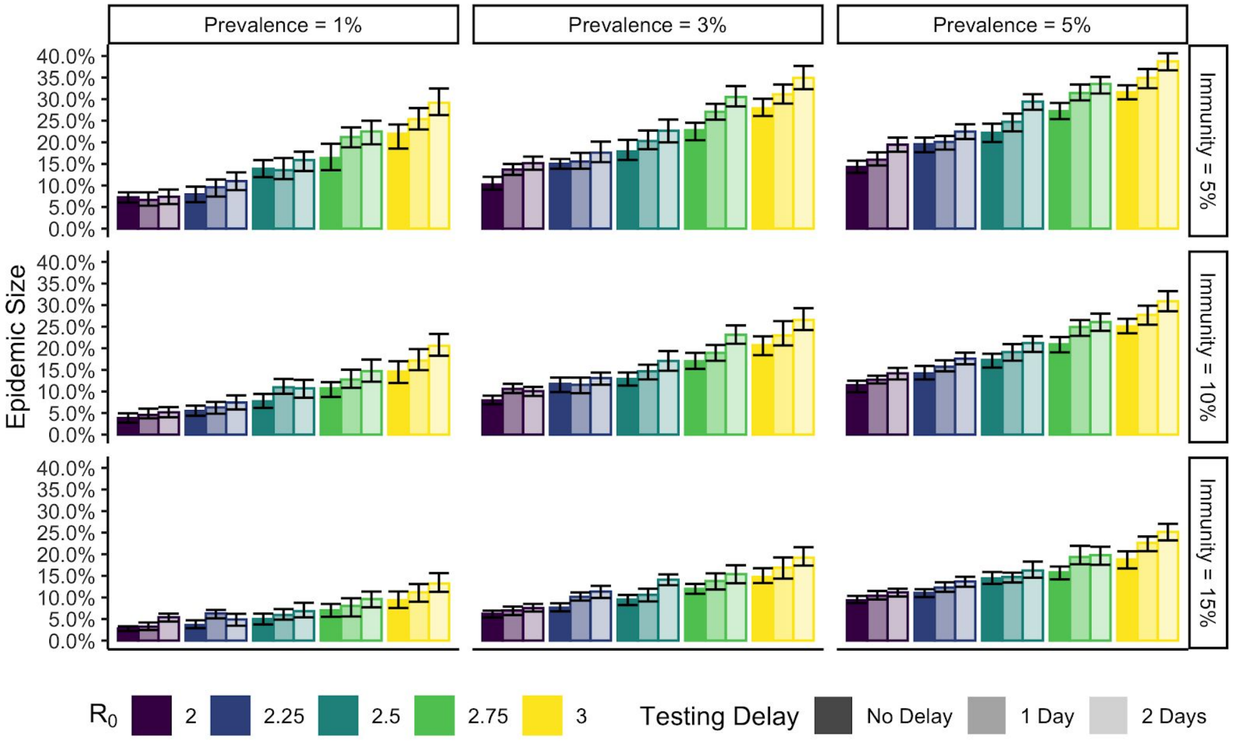

**SI Figure 2:** Epidemic size across varying levels of initial prevalence (1-5%) and initial immunity (5-15%) with various testing delays (“no delay”, “1 day”, and “2 days”) and $R_0$ (2-3). This simulation used the constant testing timeline with 2000 tests per day (10% of the population), compliance and care-seeking of 100%, 0.5 symptomatic rate, PCR sensitivity and specificity of 99%, LAMP sensitivity of 77.1%, LAMP specificity of 98%, one community introduction per 10 days to both on- and off-campus, no contact tracing, no pooling, and a 150 day semester with 50 simulations per combination.

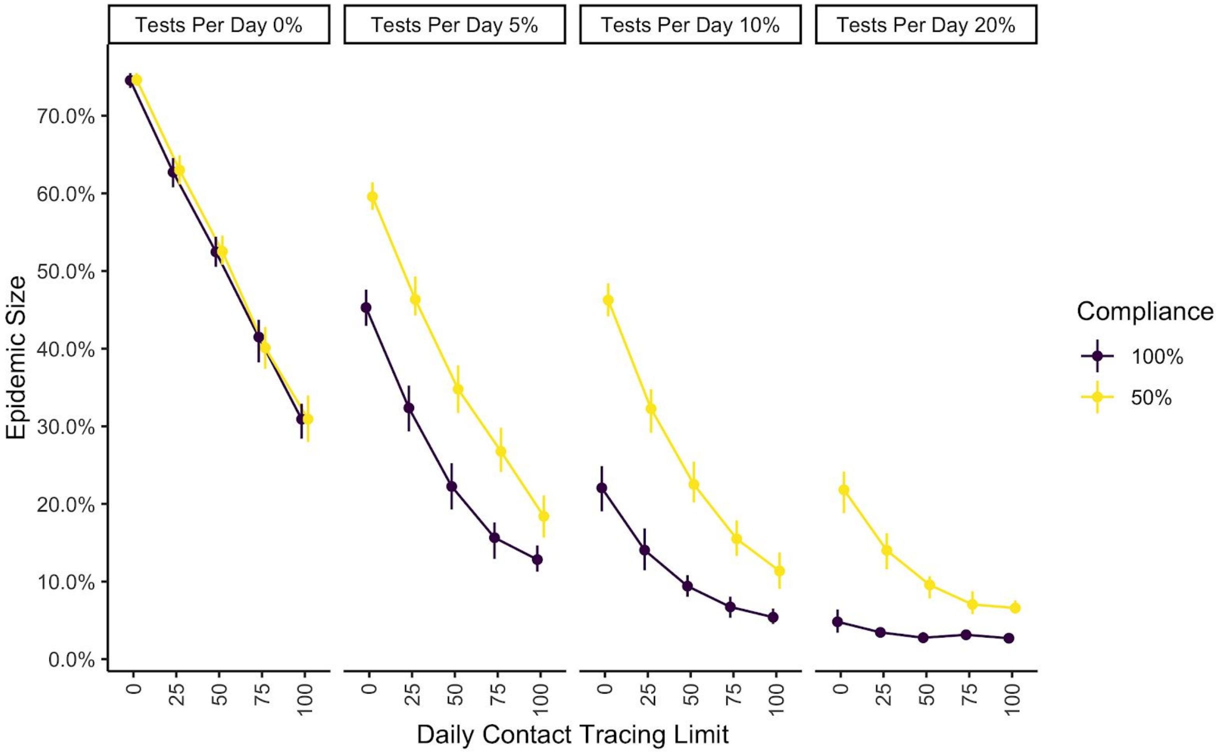

**SI Figure 3:** Epidemic size as a result of varying degrees of contact tracing effort (0 to 100 traces per day) across high (100%) and low compliance (50%) of students with testing and varying degrees of average screening tests per day (0-20%). This simulation used the constant testing timeline, with 100% care-seeking, 1% initial prevalence, 5% immunity, 0.5 symptomatic rate, $R_0$ of 3, PCR sensitivity and specificity of 99%, LAMP sensitivity of 77.1%, LAMP specificity of 98%, one community introduction per 10 days to both on- and off-campus, no contact tracing, no pooling, and a 150 day semester with 50 simulations per combination.

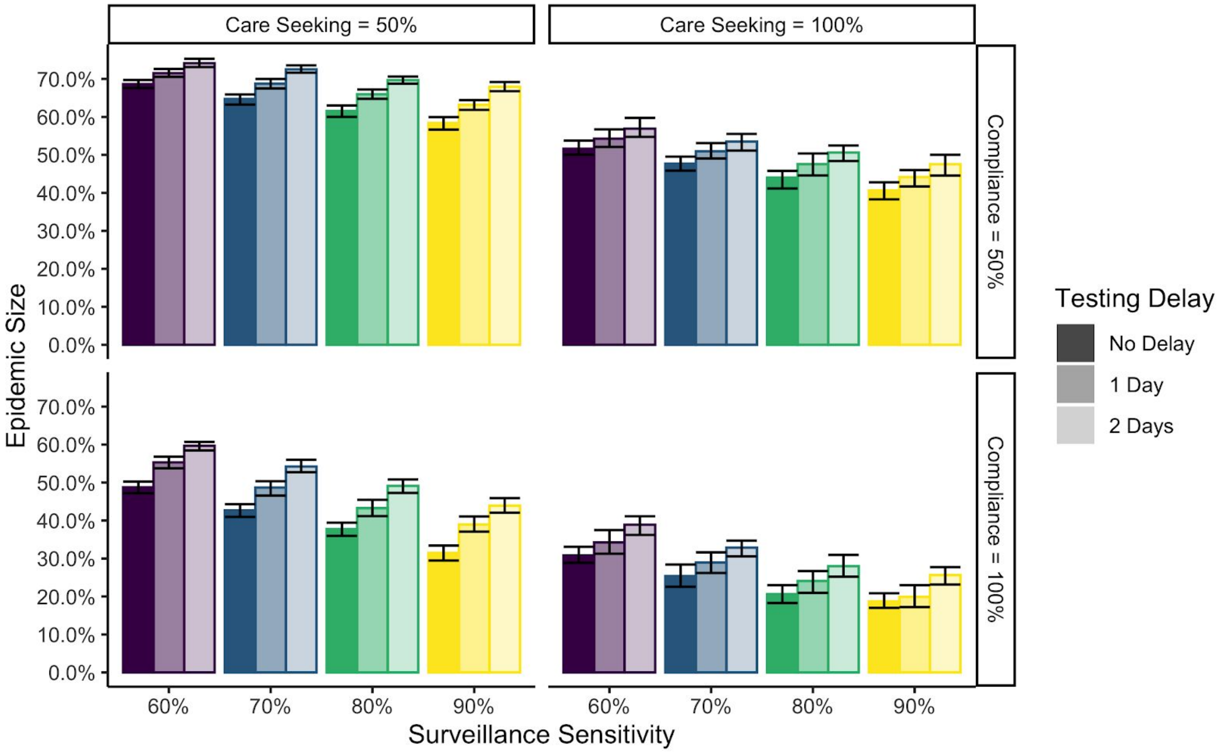

**SI Figure 4:** Epidemic size across high (100%) and low (50%) compliance and care-seeking along with various testing delays (“no delay”, “1 day”, and “2 day”) and screening test sensitivity (60-90%). This simulation used the constant testing timeline with 2000 tests per day (10% of the population), 1% initial prevalence, 5% immunity, 0.5 symptomatic rate, $R_0$ of 3, PCR sensitivity and specificity of 99%, LAMP specificity of 98%, one community introduction per 10 days to both on- and off-campus, no contact tracing, no pooling, and a 150 day semester with 100 simulations per combination.

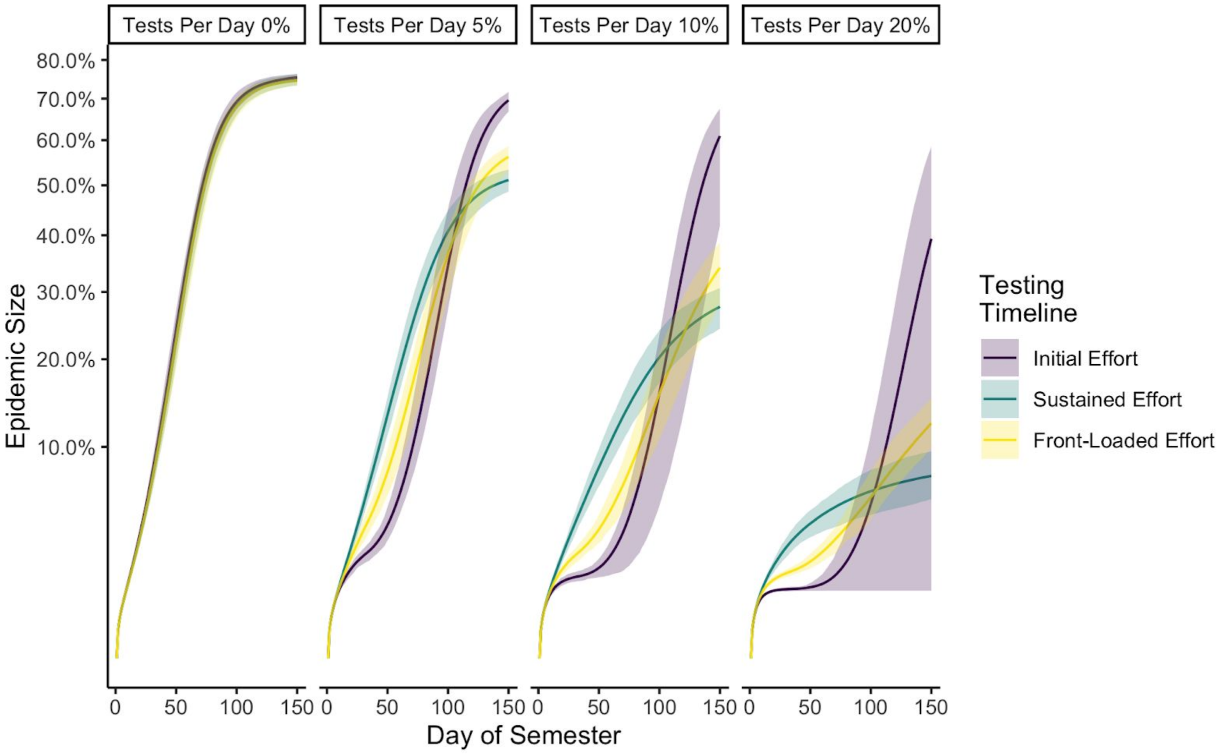

**SI Figure 5:** Epidemic size (square root-y axis) across varying testing strategies (“initial”, “sustained”, and “front-loaded” testing efforts) and testing levels (0-20%). This simulation used the constant testing timeline with 2000 tests per day (10% of the population), compliance and care-seeking of 100%, 0.5 symptomatic rate, PCR sensitivity and specificity of 99%, LAMP sensitivity of 77.1%, LAMP specificity of 98%, one community introduction per 10 days to both on- and off-campus, no contact tracing, no pooling, and a 150 day semester with 50 simulations per combination.

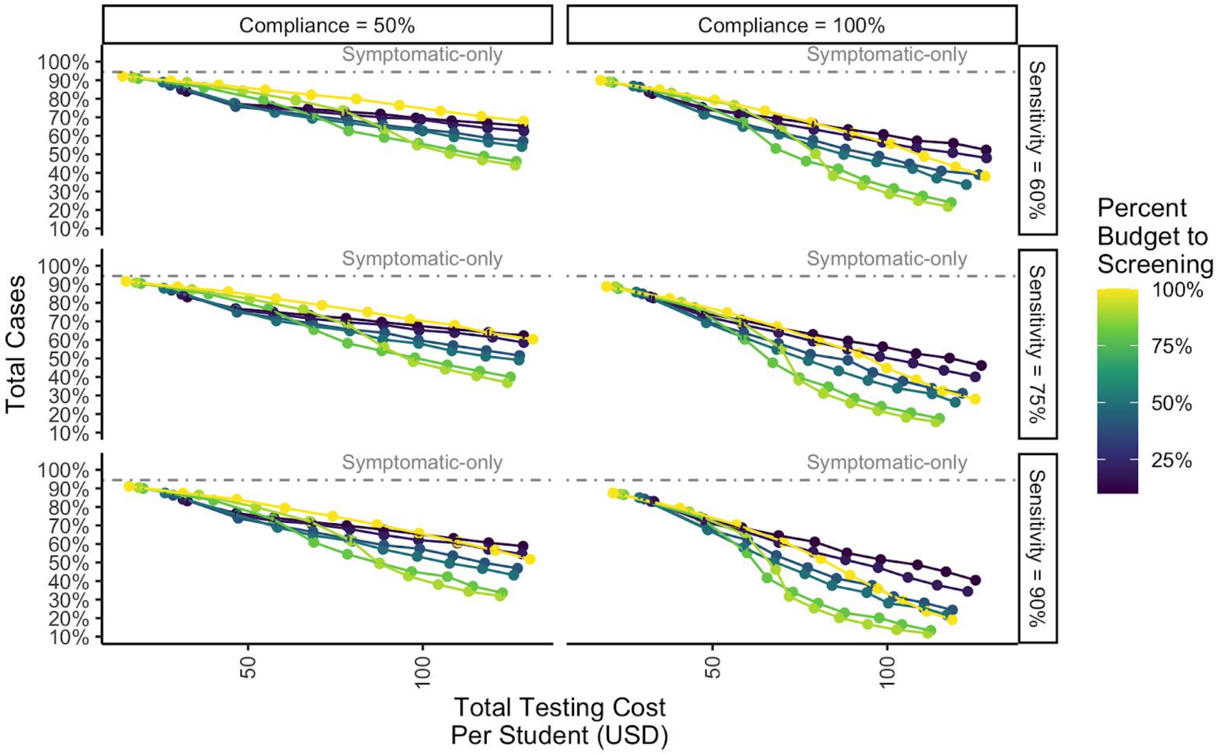

**SI Figure 6:** Epidemic size (y-axis) and total cost (x-axis; inclusive of testing and isolation costs) as they vary across levels of compliance (50-100%), sensitivity (60-90%), and the percent of testing budget devoted to screening versus diagnostic testing (0-100%) across assumed testing investments of $0 to $2 million USD. This simulation used the constant testing timeline, with 100% care-seeking, 1% initial prevalence, 5% immunity, 0.5 symptomatic rate, $R_0$ of 3, PCR sensitivity and specificity of 99%, LAMP specificity of 98%, one community introduction per 10 days to both on- and off-campus, no contact tracing, no pooling, and a 150 day semester with 25 simulations per combination. We assumed that LAMP tests were 3.5 USD, PCR tests were 12.5 USD, and that the cost to isolate on-campus students was 25 USD.

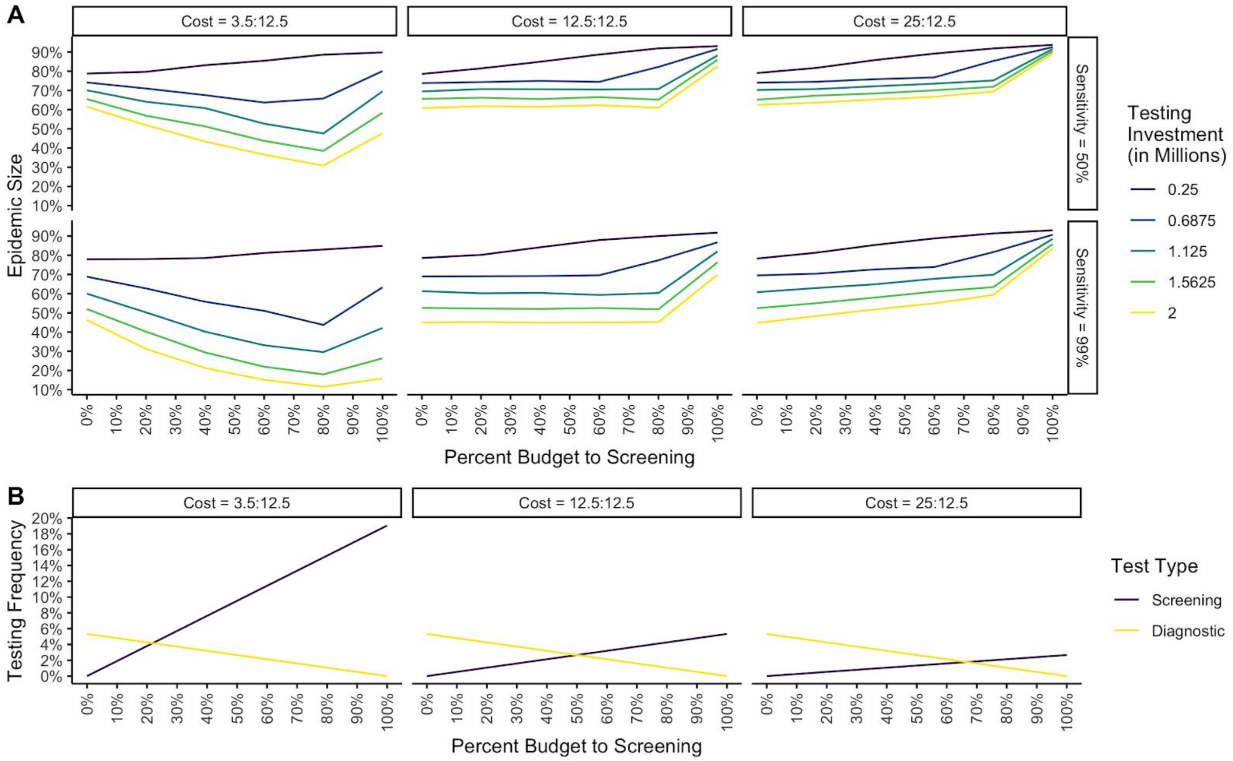

**SI Figure 7:** (A) Epidemic size (y-axis) and percent of testing investment devoted to screening (x-axis) as they vary across levels of cost ratios of the screening test to the cost of the diagnostic test (3.5:12.5 USD, 12.5:12.5 USD, 25:12.5 USD), sensitivity (50-98%), and across assumed testing investments of 0.25 to 2 million USD. Additionally, (B) the frequency of testing based on an initial investment of 2 million USD with varying cost ratios between screening and diagnostic tests with varying levels of budget devotion to screening testing. This simulation used the constant testing timeline, with 100% compliance and care-seeking, 1% initial prevalence, 5% immunity, 0.5 symptomatic rate, $R_0$ of 3, PCR sensitivity and specificity of 99%, LAMP specificity of 98%, one community introduction per 10 days to both on- and off-campus, no contact tracing, no pooling, and a 150 day semester with 25 simulations per combination. We assumed that LAMP tests were 3.5 USD, PCR tests were 12.5 USD, and that the cost to isolate on-campus students was 25 USD.

### Appendix 4: Description of university testing programs in the US

In March 2020, as cases of COVID-19 increased, most universities and residential colleges in the United States cancelled in-person classes, sent resident students home, and finished the semester online. As the fall semester approached, after a summer marked by rising numbers of COVID-19 cases, many of those same universities opted to reopen, prompting influxes of students from across the country. This migration of students to residential campuses posed a unique challenge to controlling SARS-CoV-2 epidemics both on campuses and in the surrounding communities. Additionally, a majority of university students are under the age of 30, increasing the likelihood that they may be asymptomatic carriers. This creates a situation where individuals can become infected but fail to quarantine, increasing transmission not only on campus, but potentially in the surrounding communities. Due to these unique challenges, many universities developed strategies to control viral spread, including having students test and/or quarantine for two weeks upon arrival to campus (similar to front-loaded scenario in our models), limiting the number of students staying on-campus, reducing in-person instruction, and implementing test, trace, and isolate programs to identify and quarantine active cases. Although some universities tested only symptomatic individuals, screening testing programs offer the best, if not the only measure to identify and isolate asymptomatic carriers of SARS-CoV-2. Numerous college campuses nationwide have implemented asymptomatic screening testing programs. We compiled the testing plans of the 74 land-grant universities in the United States (excluding those institutions in associated US territories such as Micronesia). Of the 72 institutions with published COVID-19 testing plans, 53 included screening testing programs. Of those institutions that implemented screening testing programs, all have opted to continue or expand their testing programs for the spring 2021 semester, indicating that these programs had significant value to the institutions that implemented them for the fall. A direct comparison of the efficacy of screening testing plans based on absolute case counts is confounded by the fact that universities with robust screening testing are likely to catch all or most of their on-campus cases, leading to higher case counts, while those institutions that test less frequently will find fewer cases. To overcome this confound, we offer the salient details of a few screening testing programs below, and clarify how these examples can be used to guide policy decision making for universities considering screening testing plans going forward.

| Land Grant Institution | Asymptomatic testing? | Fall 2020 Testing Program Details | Spring 2021 testing plan |
|---|---|---|---|
| University of Arkansas-Fayetteville | No | Symptomatic PCR testing | TBD |
| University of Arkansas-Pine Bluff | No | Symptomatic PCR testing | TBD |
| University of Idaho | No | Symptomatic PCR testing | Implementing surveillance testing program, details TBD |
| White Earth Tribal and Community College | No | Mostly online classes | TBD |
| Leech Lake Tribal College | No | Mostly online classes | TBD |
| Alcorn State University | No | Symptomatic PCR testing | TBD |
| Montana State University | No | Symptomatic PCR testing | Implementing surveillance testing program for residential students at the beginning of the semester |
| University of Nevada Reno | No | Symptomatic PCR testing | TBD |
| North Carolina A and T State University | No | Symptomatic PCR testing | TBD |
| Oklahoma State University | No | Symptomatic PCR testing | TBD |
| Langston University | No | None indicated | TBD |
| South Carolina State University | No | One special testing event was held, most courses held online | Mostly virtual delivery and limited housing available |
| Virginia State University | No | No published plan | TBD |
| University of Missouri | No | Symptomatic PCR testing | TBD |
| University of Alaska Fairbanks | No | Symptomatic PCR testing and arrival testing for students and employees arriving from out of state or living in residence halls. | TBD |
| Central State University | No | Symptomatic testing, and arrival testing | TBD |
| Kentucky State University | Yes | Periodic testing events throughout the semester | TBD |
| Fond du Lac Tribal and Community College | Yes | Periodic testing events throughout the semester | TBD |
| Mississippi State | Yes | Periodic testing events throughout | TBD |

| | | |
|---|---|---|
| University | | the semester |

| | | | |
|---|---|---|---|
| Alabama A and M University | Yes | Initial testing of all students returning to campus and sentinel testing (2.5% of random student population) during the semester | Remote learning for the spring |
| Auburn University | Yes | Opt-in sentinel population (2.5-5% of community) testing | Continuing, potentially expanding current surveillance strategy, details TBD |
| Tuskegee University | Yes | Initial entry test for all students, sentinel testing (15% of student body) throughout the semester | TBD |
| University of Arizona | Yes | Random PCR tests of asymptomatic students and staff | Continuing, potentially expanding current surveillance strategy, details TBD |
| University of California-Berkeley | Yes | Required for resident students 2x/week and recommended/available to all | Continuing current surveillance strategy and flexible course delivery options |
| Colorado State University | Yes | Saliva-based screening freely available to students, faculty and staff | Continuing current surveillance strategy and flexible course delivery options |
| University of Connecticut | Yes | Entry and random surveillance testing required for all students | In person following an initial 2 week online period so all students can quarantine |
| University of Delaware | Yes | Random surveillance testing | Continuing current testing program |
| Delaware State University | Yes | Twice weekly for all dorm residents and staff and students receiving in person classes; most courses delivered online | Continuing current testing program |
| University of Florida | Yes | Testing for symptomatic students, those working in the clinic, and arriving from specific states | Expanding testing to all undergraduates |
| Florida A and M University | Yes | Sentinel testing of both students and faculty | TBD |
| University of Georgia | Yes | 500+ tests a day for students, staff and faculty | Continuing current surveillance strategy and flexible course delivery options |
| Fort Valley State University | Yes | Mandatory random testing of 5% of students residing on campus | |
| University of Hawaii | Yes | Surge testing, large scale testing performed at different points | Continuing current surveillance strategy |

| | | | |
|---|---|---|---|
| | | during the semester | |
| University of Illinois | Yes | Mandatory twice a week testing for all students | Continuing current surveillance strategy and flexible course delivery options |
| Purdue University | Yes | Random testing throughout semester for students, faculty, and staff | Continuing current surveillance strategy and flexible course delivery options |
| Iowa State University | Yes | On campus testing for students, staff and faculty | Continuing current surveillance strategy and flexible course delivery options |
| Kansas State University | Yes | Voluntary asymptomatic testing for students, staff and faculty, expanded throughout the semester | TBD |
| University of Kentucky | Yes | Arrival testing followed by random surveillance | TBD |
| Louisiana State University | Yes | Free testing available to anyone on campus, but participation is not required | Continuing current surveillance strategy and flexible course delivery options |

| | | | |
|---|---|---|---|
| Southern University and A and M College | Yes | Free testing available to anyone on campus, but participation is not required | Continuing current surveillance strategy and flexible course delivery options |
| University of Maine | Yes | Arrival testing followed by random surveillance | Continuing current surveillance strategy and flexible course delivery options |
| University of Maryland College Park | Yes | Arrival testing followed by required monthly testing for all students that are on campus | Continuing current surveillance strategy and flexible course delivery options |
| University of Massachusetts Amherst | Yes | Weekly tests required for all students | Continuing current surveillance strategy with a majority of classes online and limited students allowed on campus |
| Massachusetts Institute of Technology | Yes | All students are tested twice a week | Continuing current surveillance strategy and flexible course delivery options |
| Michigan State University | Yes | Free testing available to anyone on campus but not required. Periodic testing of a volunteer sentinel population throughout semester | Additional on campus classes and expanded testing with required registration in surveillance programs for on campus students and undergrads |

| | | | |
|---|---|---|---|
| University of Minnesota | Yes | Single saliva-based test for each student | TBD |
| Lincoln University | Yes | Random testing throughout semester for both students, faculty and staff | Starting remote in Jan., with plan to transition to in-person classes in March |
| University of Nebraska Lincoln | Yes | Free testing available to anyone on campus but participation is not required | TBD |
| University of New Hampshire | Yes | Required testing of all students twice a week | Continuing current surveillance strategy and flexible course delivery options |
| Rutgers, the State University of NJ | Yes | Available on request based on risk assessment for the limited number of individuals currently on campus | Expanding on campus activities, testing program TBD |
| New Mexico State University | Yes | Required arrival testing, and weekly surveillance testing of athletes and 25% of student population | Continuing current surveillance strategy and flexible course delivery options |
| Cornell University | Yes | Testing of all students twice per week | Continuing current surveillance strategy and two negative tests mandatory to return to campus |
| North Carolina State University | Yes | Limited voluntary surveillance testing | Continuing current surveillance strategy, and requiring a negative test to return to campus |
| North Dakota State University | Yes | Free asymptomatic testing started for students at a few scheduled testing events | Expanding current program to regular voluntary surveillance testing of all students |
| Ohio State University | Yes | Free testing available to anyone on campus but participation is not required | TBD |
| Oregon State University | Yes | Available to a subset of students on a limited basis | Expanding to all faculty, staff and students voluntarily and requiring resident students' participation |
| Pennsylvania State University | Yes | Daily tests of ~1% of the population | Arrival testing followed by continuing or expanding existing program |
| University of Rhode Island | Yes | Mandatory surveillance testing of all students throughout the semester, staff participation is voluntary | Continuing current surveillance strategy with expansion TBD and flexible course delivery options |
| Clemson University | Yes | Already updated for spring, unknown fall status | Required routine surveillance testing |
| South Dakota State | Yes | Students are eligible, voluntary | Continuing current surveillance |

| | | | |
|---|---|---|---|
| University | | | strategy |
| University of Tennessee | Yes | Testing available for all asymptomatic students and staff; required saliva pooled testing for campus residents | TBD |
| Tennessee State University | Yes | Free testing available to anyone on campus but participation is not required | TBD, 85% remote learning |
| Texas A and M University | Yes | Free testing available to anyone on campus and general public but participation is not required | Continuing current surveillance strategy and flexible course delivery options |
| Prairie View A and M University | Yes | Free testing available to anyone on campus but participation is not required | TBD |
| Utah State University | Yes | Free testing available to anyone on campus but participation is not required | TBD |

| | | | |
|---|---|---|---|
| University of Vermont | Yes | All students are tested weekly | Continuing current surveillance strategy and flexible course delivery options, with possible return testing required |
| Virginia Tech | Yes | Free testing available to anyone on campus but participation is not required | Required arrival testing, surveillance not yet announced |
| Washington State University | Yes | Asymptomatic testing is available to anyone on campus, course delivery is primarily virtual | Mostly virtual delivery, continuing current surveillance and increasing all around testing capacity for all |
| West Virginia University | Yes | Goal to test 200 people/week with a saliva based assay | Continuing current surveillance strategy and flexible course delivery options |
| West Virginia State University | Yes | Random tests of on- campus individuals | Continuing current surveillance strategy and flexible course delivery options, with testing required for return to campus |
| University of Wisconsin-Madison | Yes | Surveillance testing available to students, faculty and staff, required in campus housing | Expanding testing program to test all students twice/week with saliva-based qPCR |
| University of Wyoming | Yes | Undergraduate students tested twice weekly with saliva-based pooled testing, follow up PCR test required | Continuing, potentially expanding current surveillance strategy, details TBD |

| | | | |
|---|---|---|---|
| | | for members of positive pools | |
| Red Lake Nation College | No Information Available | No Information Available | No Information Available |
